# Supervised Learning for CT Denoising and Deconvolution Without High-Resolution Reference Images

**DOI:** 10.1101/2023.08.31.23294861

**Authors:** Andrew D. Missert, Scott S. Hsieh, Andrea Ferrero, Cynthia H. McCollough

## Abstract

**Purpose:** Convolutional neural networks (CNNs) have been proposed for super-resolution in CT, but training of CNNs requires high-resolution reference data. Higher spatial resolution can also be achieved using deconvolution, but conventional deconvolution approaches amplify noise. We develop a CNN that mitigates increasing noise and that does not require higher-resolution reference images.

**Methods:** Our model includes a noise reduction CNN and a deconvolution CNN that are separately trained. The noise reduction CNN is a U-Net, similar to other noise reduction CNNs found in the literature. The deconvolution CNN uses an autoencoder, where the decoder is fixed and provided as a hyperparameter that represents the system point spread function. The encoder is trained to provide a deconvolution that does not amplify noise. Ringing can occur from deconvolution but is controlled with a difference of gradients loss function term. Our technique was demonstrated on a variety of patient images and on ex vivo kidney stones.

**Results:** The noise reduction and deconvolution CNNs produced visually sharper images at low noise. In ex vivo mixed kidney stones, better visual delineation of the kidney stone components could be seen.

**Conclusions:** A noise reduction and deconvolution CNN improves spatial resolution and reduces noise without requiring higher-resolution reference images.

## 1 Introduction

Spatial resolution is an important image quality metric for every medical imaging modality, including computed tomography (CT). Tasks such as lung nodule characterization, bone fracture detection, and kidney stone assessment all benefit from higher spatial resolution. Historically, in-plane spatial resolution has improved slowly, increasing from 10 line pairs per cm in 1980 to about 20 line pairs per cm in 2015 (1). Other modalities have witnessed much faster progress: MRI has seen spatial resolution improvements with every magnet strength upgrade; ultrasound has seen spatial resolution improvements from higher frequency transducers enabled by lower noise electronics, and PET scanners have seen spatial resolution improvements from time-of-flight coincidence detectors. The primary reason for slower progress in CT is the nature of the pixelated scintillator-photodiode detector, in which smaller pixels reduce detection efficiency. When these limitations are overcome using, for example, recent photon counting detectors that increase fill factor (2, 3), there is still the more fundamental limitation of the noise power spectrum: higher spatial frequencies are always associated with increased noise because they are sampled by fewer views (4).

Absent new hardware, software approaches can be employed to improve the resolution of a reconstructed CT image. We consider two categories of software approaches that enhance the representation of small-scale image features: super-resolution and deconvolution. We define super-resolution as a technique that improves the sampling rate of the image, e.g. increasing the matrix size from 512×512 pixels to 1024×1024 pixels. We define deconvolution as a technique that inverts the blurring caused by the finite spatial resolution of the imaging system. In photographic images acquired with a wide depth of field and sharp focus, super-resolution is often the best approach for enhancing fine detail. In CT images, the point spread function is often larger than the reconstructed voxel size in high-resolution protocols, so deconvolution approaches would improve spatial resolution even without increasing the matrix size.

Although analytic approaches exist for both deconvolution and super-resolution, they are of limited practical use due to the ill-posed nature of the problem in the presence of quantum noise. Traditional approaches to deconvolution employ some type of regularization to increase robustness to noise. Recent studies have demonstrated that deep learning models such as convolutional neural networks (CNNs) can be used. These nonlinear CNNs encode prior information from training examples that can enhance the accuracy of the output high-resolution images. These CNNs have been demonstrated very effectively for photographic images (5-7), and progress has also been reported for CT images (8, 9). Two of the most significant challenges for applying these techniques to CT imaging are: 1) the high amount of noise in CT compared to conventional photography, and 2) the lack of a high-resolution ground truth reference needed to optimize the CNN parameters.

Previous groups using CNNs for CT super-resolution have reported the ability to recover standard-resolution CT from downsampled, low-resolution CT, but they have not convincingly demonstrated the ability to increase the resolution of standard-resolution CT. For example, Park et al. trained a CNN to map the average of a stack of adjacent axial slices to the central slice (8). Umehara et al. trained a CNN to map from a 2x downsampled chest CT image to an original-resolution chest CT image, outperforming simpler schemes such as linear or bicubic interpolation (10). You et al. used generative adversarial networks with several constrains, including cycle consistency, but also required high-resolution CT images during training (9). It is not established that these techniques, demonstrated for the task of mapping half-resolution to standard-resolution, could be applied without modification to map from standard-resolution to double-resolution. While such a leapfrog approach has been applied successfully in the context of noise reduction, there is reason to believe it would be less effective for super-resolution. The texture of noise and the scale of the point spread function are not invariant across resolution scales. For this reason, noise reduction CNNs do not generalize well across reconstruction kernel changes (11, 12).

The purpose of this work is to develop a noise reduction and resolution enhancement CNN that does not require high-resolution reference images. We will use a novel autoencoder scheme and provide the network with an estimate of the point spread function, which the CNN will use to perform resolution enhancement. We will apply the CNN to a variety of patient datasets and to *ex vivo* kidney stones. Kidney stones are a special application because several kinds of kidney stones appear similar in CT imaging, but knowing the composition of a kidney stone is a cornerstone of the management of recurrent nephrolithiasis (13). Today, stones are collected after passing naturally or when extracted surgically and are analyzed chemically to determine chemical composition, but a method to determine stone composition from CT images would be much more convenient. Today, dual energy CT can reliably differentiate uric acid from nonuric acid stones (14), but differentiation of mixed stones or of nonuric acid subtypes can only occur under ideal conditions (15). Higher resolution CT could be useful for this task.

## 2 Materials and Methods

### 2.1 General design and motivation

Let the overall effect of the finite spatial resolution of the imaging system be modeled as a function K : Y → *X* that maps from a high-resolution image representation *y* ∈ *Y* to a corresponding low-resolution image *x* ∈ *X*. Solving the image deconvolution problem is equivalent to finding a solution Φ_deconv_ that is the inverse of *K*. Specifically, we need Φ_deconv_ to satisfy the following condition:

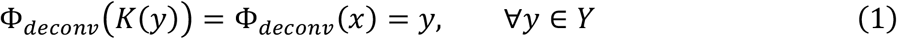

One major challenge to solving Eq. 1 is noise. Noise arises from x-ray quantum statistics and may further be exacerbated by electronic noise in the detector. In the presence of noise, we can abstractly decompose the observed CT image z as *z* = *K*(*y*) + δ, where δ denotes the image features arising from random noise. The expression from Equation 1 then becomes:

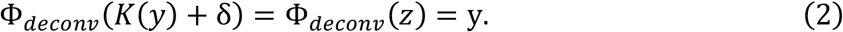

We can reduce expression in Equation 2 by introducing a noise-reduction operator Φ_*denoise*_that satisfies the condition:

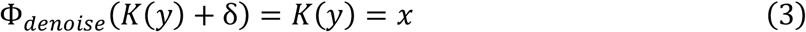

When this operator is composed with Φ_*deconv*_, Equation 2 reduces to the form of Equation 1:

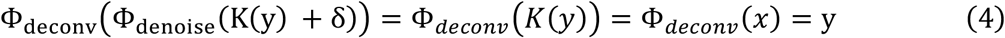

Defining explicit models for the operators Φ_*deconv*,_ and Φ_*denoise*_is difficult in practice. Considering that noise δhas some finite chance of taking on any possible image, perfect solutions to Φ_*deconv*,_ and Φ_*denoise*_cannot exist. Instead, they can only be optimized or trained to be effective with typical noise realizations. In this work, we chose to model these operators as deep CNNs with tunable parameters θ that are optimized using supervised learning. Through the parameter optimization process, we aim to find parameters θ^*^ such that:

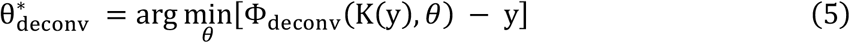

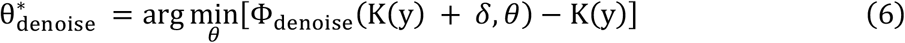

The resulting CNN models Φ_*deconv*,_(*x*, θ *) and Φ_*denoise*_(*x*, θ *) are then approximate solutions to the deconvolution problem posed by Equation 4.

To evaluate the expression in Equation 5, the high-resolution image *y* must be known. This is problematic in a practical setting, since the CT scanner only produces images with limited spatial resolution, corresponding to *K*(*y*). The corresponding high-resolution image *y* is unknown. We circumvent this problem by applying the blurring operator *K* to both sides of Equation 4. This results in the modified optimization condition:

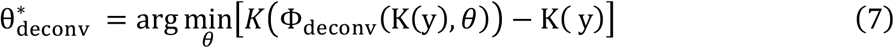

which is also minimized by the desired solution Φ_*deconv*,_(*K*(*y*), θ) = *y*. Equation 7 is critical to the optimization process in this work, since it allows us to optimize the CNN models using supervised learning without explicit knowledge of the high-resolution ground-truth *y*. The data preparation process and the optimization configurations for Φ_*deconv*,_(*x*, θ) and Φ_*denoise*_(*x*, θ) are described in greater detail in the following sections.

### 2.2 Denoising model optimization

We first optimized the Φ_*denoise*_(*x*, θ) operator, implemented as a CNN. The architecture of this CNN resembles the U-Net design that is commonly used for tasks related to medical image segmentation (16), but some modifications were made.

First, a preprocessing layer that applies common CT window settings to the input values, and rescales the outputs to the range [0,1], was used. The outputs for each window level, along with the rescaled full-range image, are stacked along the channel dimension before being passed to the convolutional layers. This allows the downstream layers to adjust the weights of different CT image features as part of the optimization process. Second, compared to the original U-Net, the overall architecture is wider (more filters per layer) and less deep (fewer downsampling steps), which reflects the localized nature of the noise reduction task compared to semantic image segmentation. A schematic of the CNN architecture and the relevant models can be found in Figure 1.

**Figure 1:**
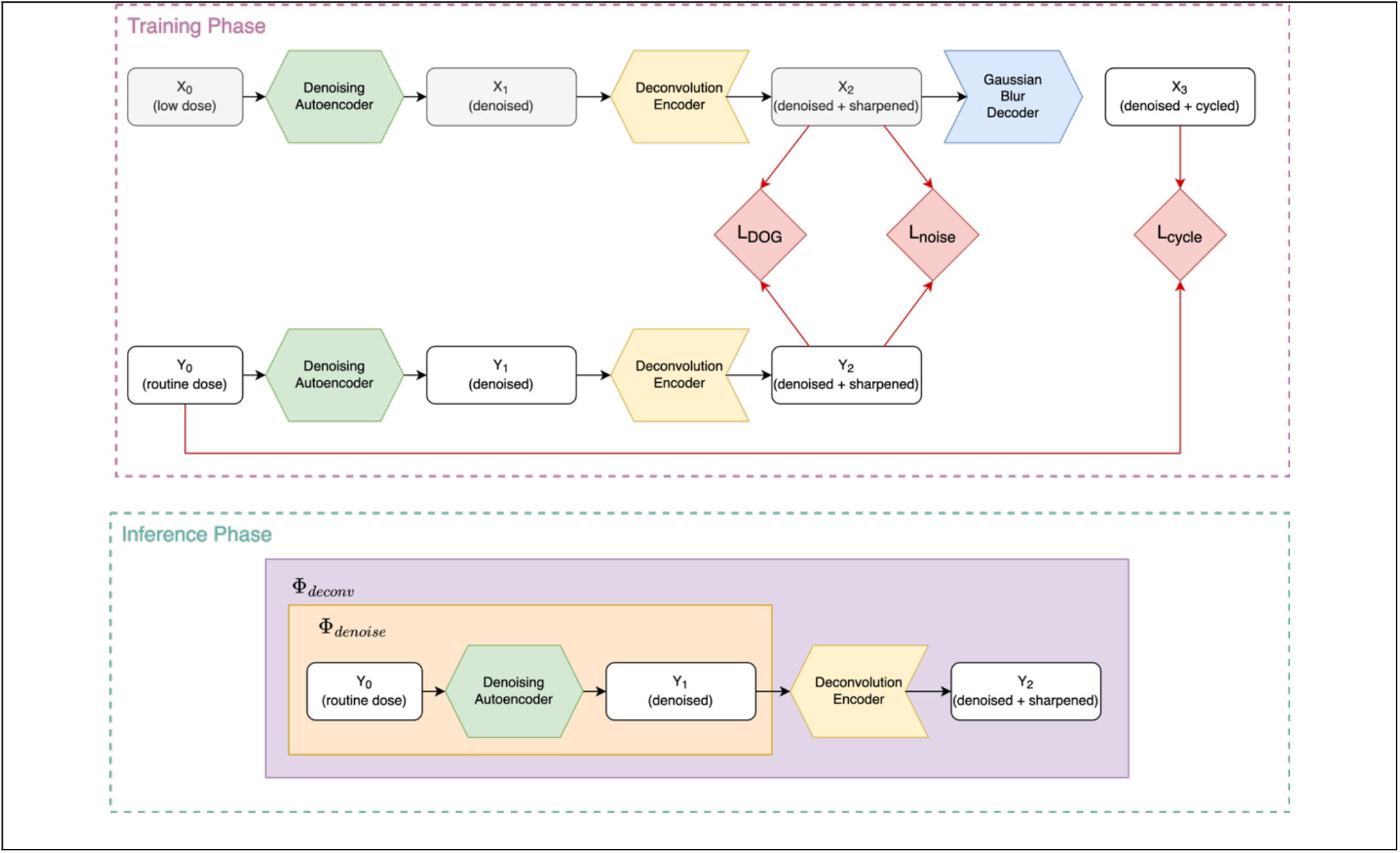
Schematic diagram of the CNN models during training and inference phases. The denoising autoencoder and the deconvolution encoder are modeled using variations of the U-Net architecture. The decoder approximates the finite spatial resolution of the imaging system using fixed convolutional layer with a Gaussian kernel. White boxes denote image-domain representations at various points in the processing chain. Red arrows connect images to the associated loss functions (red boxes) that are optimized during the training phase. The grey boxes denote the final output images during the inference phase.

Training data for optimizing the model using a bootstrapping approach. Image patches with a shape of 64×64×7 pixels were extracted from the original CT images, and 10 independent noise realizations were added using a noise insertion tool (17). A total of 150,000 images patches generated from 9 of the 10 noise simulations was used for training. From the remaining noise simulation, 200 patches were extracted as validation data to monitor for overfitting during the optimization process.

During the optimization phase, the simulated high-noise image patches were used as inputs to the CNN model, and the corresponding original images were used as target outputs. Including multiple independent noise realizations as inputs acts as a form of data augmentation to reduce overfitting and encourage the model output to be invariant with respect to the addition of typical CT noise features. The model was optimized to minimize the mean squared difference between the output images and original (low-noise) target images for 50 epochs using the Adam optimizer with a maximum learning rate of 0.001 and a step decay factor of 0.25 every 16 epochs.

### 2.3 Deconvolution model and optimization

The deconvolution operator Φ_*deconv*,_(*x*, θ) was also modeled as a CNN with a U-Net-based architecture, illustrated in Figure 1. The operator *K*(*x*) that represents the finite spatial resolution of the imaging system was modeled using a convolution layer with a Gaussian kernel at fixed width of 0.23 mm. The width of the Gaussian kernel was empirically tuned. The dataset used for optimizing the deconvolution model consisted of paired high-noise (*x*) and low-noise (*t*) image patches as described previously.

During the training phase, the simulated high-noise image patches *x* were first passed through a CNN denoising autoencoder that was pre-trained according to Section 2.2. The resulting low-noise images *x*_*denoised*_ were then used as inputs to the deconvolution model Φ_*deconv*,_ to produce an output “encoded” representation *x*_*encoded*_. The convolutional blurring operator *K* was then applied to the encoded image, to produce an output decoded image *x*_*decoded*_.

Optimization of Φ_*deconv*,_ was driven by a minimization of a loss function consisting of three terms. The first term was a standard L2 cycle consistency term of the form:

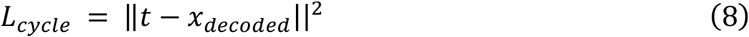

This term enforced consistency between t and *x*_*decoded*_= *K*(Φ_*deconv*,_(*x*)), which pushes Φ_*deconv*,_ to approximate the desired inverse condition in Equation 7. The second term was a noise-invariance term of the form:

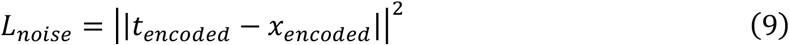

which enforces consistency between the encoded (deconvolved) representations of the low-noise and high noise images and further reduces noise in the encoded domain. Finally, a “direction-of-gradient” (DOG) loss term of the form:

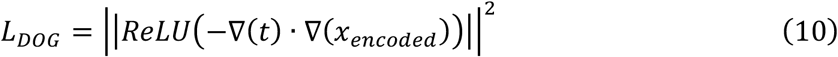

was implemented to reduce the presence of high frequency “ringing” artifacts. This term is minimized when the gradients of the encoded image and the reference image are not mis-aligned. The loss functions from the weighted sum of Equations (8-10) was minimized over 50 epochs of gradient descent with the Adam optimizer with a maximum learning rate of 0.001 and a step decay factor of 0.25 every 16 epochs.

## 3 Results

Figure 2 illustrates the trained autoencoder without noise invariance and DOG regularization. This sharpens the image but also introduces ringing. Ringing is a more general phenomenon that is seen in analytic CT reconstruction with certain sharp kernels, where edge overshoot is considered an acceptable tradeoff for improved visualization. Careful tuning of the regularization weights reduces these ringing artifacts.

**Figure 2.**
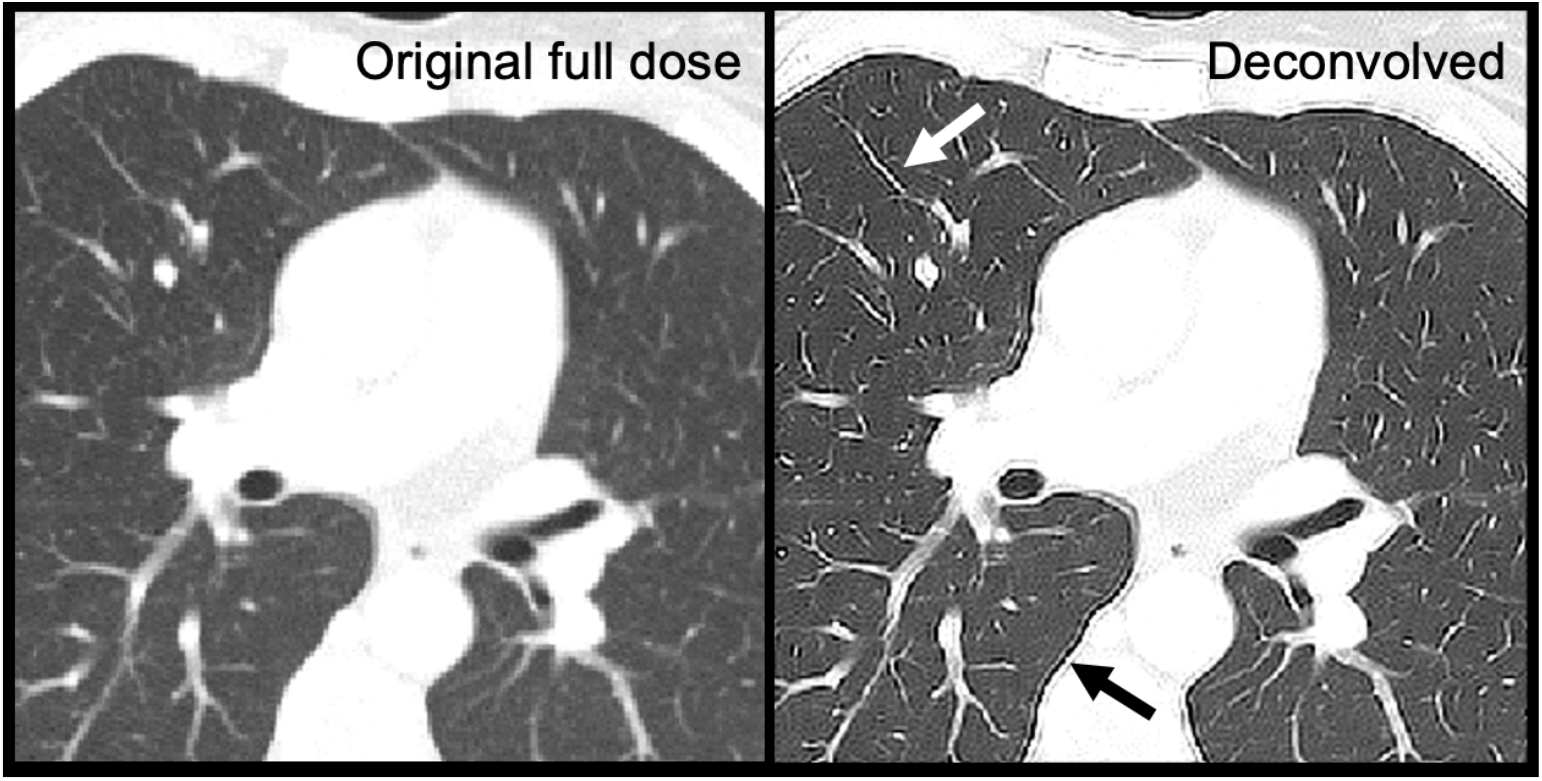
Autoencoder trained without additional regularization. On the right panel, the white arrow points to the enhanced sharpening of the airways, while the black arrow points to ringing artifacts that occur around the mediastinum.

Figure 3 shows the application of our denoising and deconvolution CNN on a routine abdominal scan. The borders of the kidneys appear to be better defined after denoising and deconvolution, and noise in the liver is greatly reduced. A ground truth image (higher resolution CT) is not available, as would be typical in most applications of super-resolution CT. Ringing is mostly controlled from regularization.

**Figure 3.**
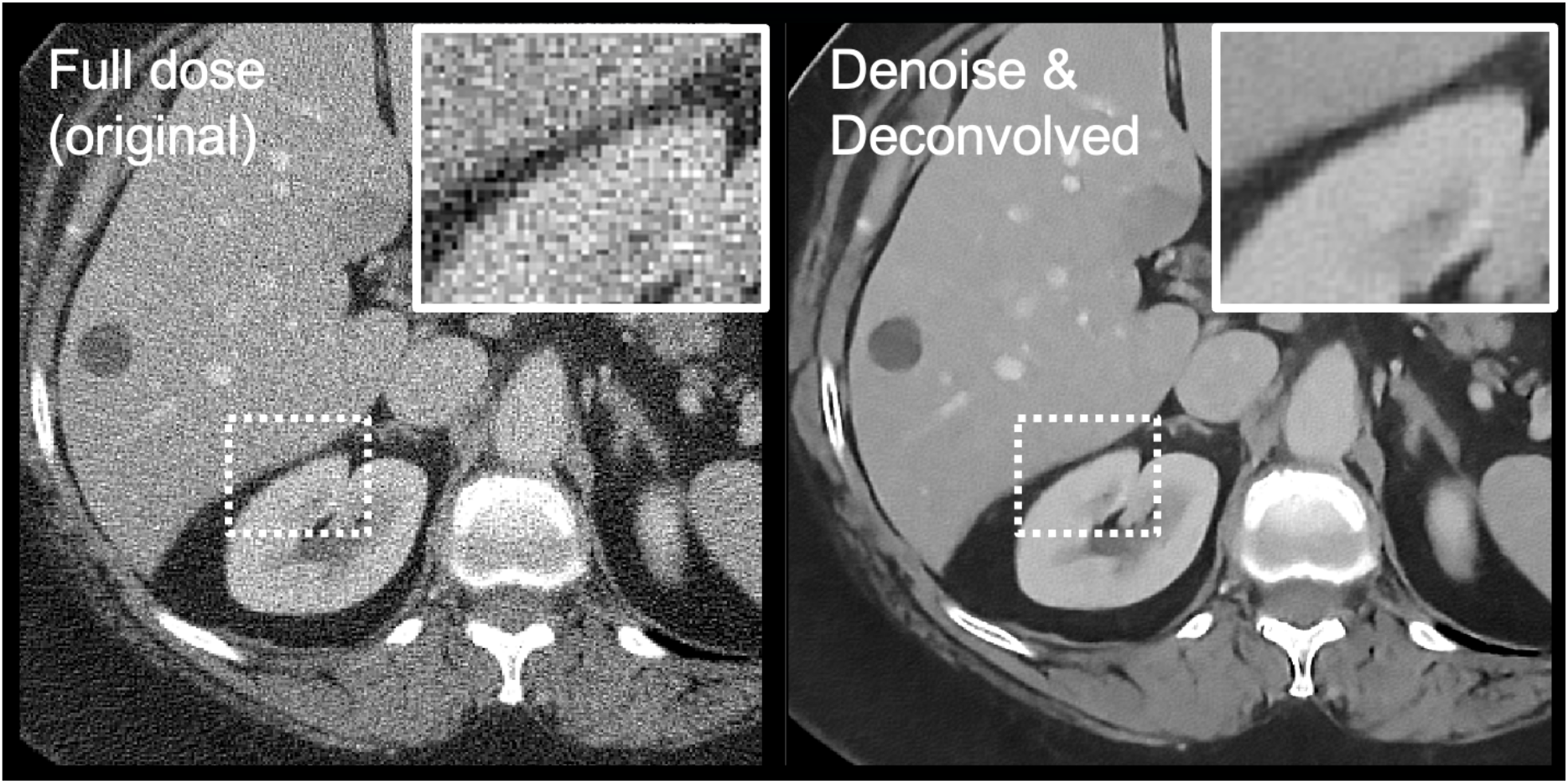
Denoising and deconvolution of a routine slice of an abdominal scan shows apparent reductions in noise and sharpening of boundaries.

Models were also applied to *ex vivo* kidney stones scanned in an ultrahigh resolution (UHR) protocol. UHR scans were acquired with a SOMATOM Force using a Uq69 kernel (the sharpest available quantitative kernel) and a CTDI of approximately 60 mGy, much higher than would be used in routine clinical practice and reconstructed to a 100 mm FOV. Figure 4 shows the kidney stones with the denoising model alone as well as the combined model. Some finer structures are more easily seen after the deconvolution step.

**Figure 4:**
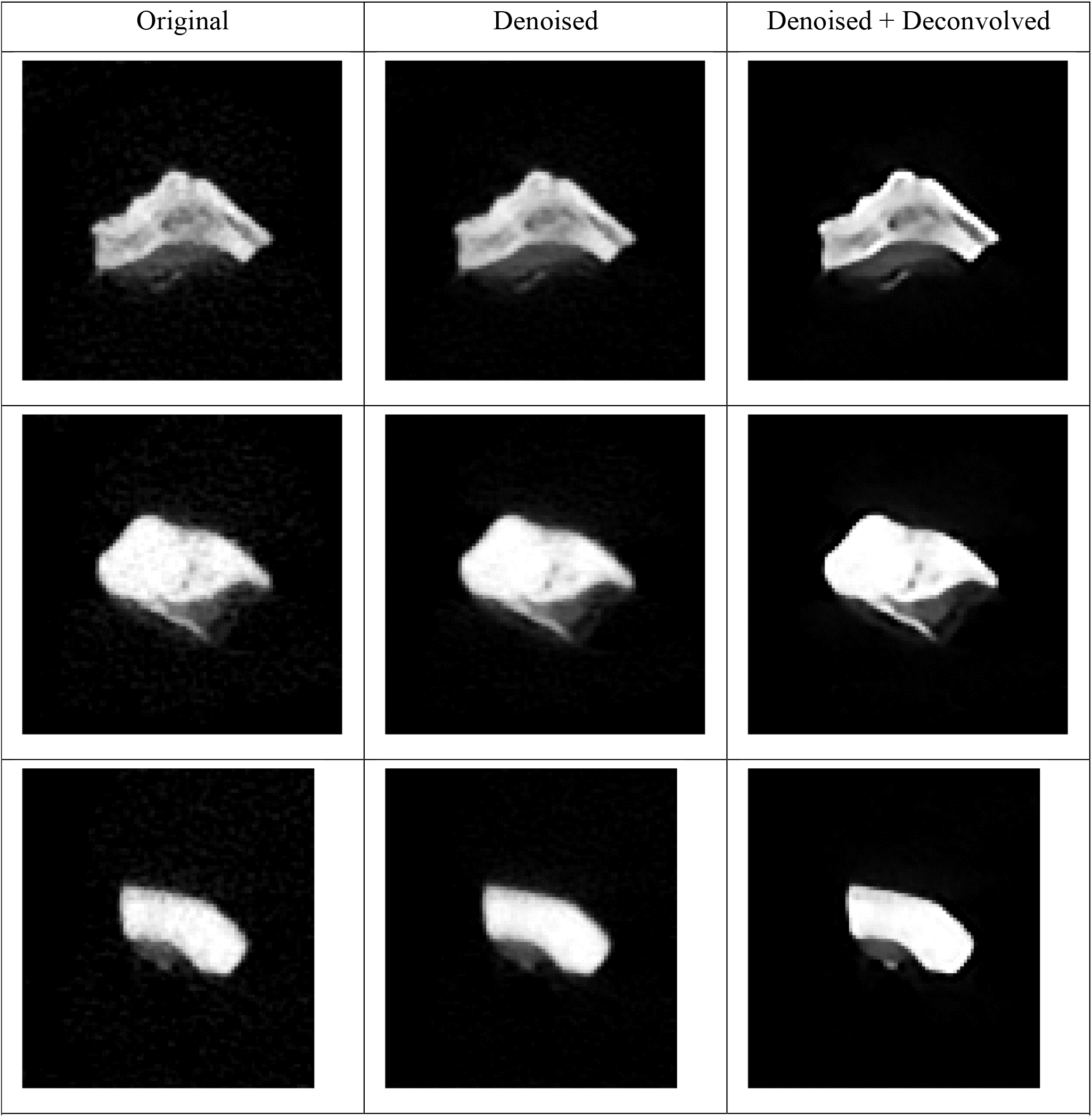
Examples of ex vivo kidney stones at various processing stages with a windowing interval of [0, 2500] HU. Left column shows the original UHR CT images. Central column shows the images after processing with the denoising model only. Right column shows the images after processing with the denoising and deconvolution algorithm.

## 4 Discussion

Supervised deep learning for super-resolution CT is challenged by the lack of high-resolution reference images. We use deconvolution to eliminate the need for high-resolution reference images and instead supply an estimated point spread function. Our results suggest that improved visualization can occur, even when the original protocol is optimized for maximum spatial resolution. Artifacts are present but are controlled by regularization. In clinical practice, we expect that the value of our approach will be task-dependent. For tasks that do not require very high spatial resolution, it may be sufficient to simply reconstruct at a sharper kernel than normal and denoise this image to control the elevated noise. We previously demonstrated that joint denoising of multiple kernels may be valuable (18). However, when it is desirable to go beyond the sharpest kernel of the CT scanner, there may be incremental value of the deconvolution autoencoder approach, as seen in Figure 4. Besides kidney stone assessment, other clinical tasks in this category include quantification of lumen diameter (19) and evaluation of the temporal bone (20).

This work is an example of the more general problem of missing training labels. In CT noise reduction, other authors have addressed this problem by using noise-to-noise mappings (21) or by using a CycleGAN approach (22). A major difference is that in noise reductions, several good options exist to create realistic training labels, including phantom-based noise insertion (23), analytic estimation and backprojection of CT noise of the correct texture (24), or insertion of very realistic noise with cooperation of the scanner vendor (17). Noise reduction CNNs created by CT vendors can be expected to have especially good modeling of the noise pipeline. On the other hand, for the super-resolution problem, realistic estimates of the training labels cannot be created, at least for patient scans and when operating a CT scanner at its maximum spatial resolution.

As the adoption of noise reduction CNNs continues to increase, our algorithm provides a way for further image enhancement in high-resolution protocols. Deep learning reconstruction and denoising products have already been demonstrated to provide some clinical benefit above iterative reconstruction (25). Many tools currently being developed provide control over denoising strength but not image sharpness. An autoencoder chained to the primary noise reduction CNN provides a mechanism of providing sharper images that could further tune image quality. Currently, tasks that require maximum spatial resolution are reconstructed with frequency boosting ultrasharp FBP kernels. Our work provides a means for translating these reconstruction algorithms into deep learning reconstruction, where additional noise reduction is possible.

## Data Availability

Data from this study are not available due to patient confidentiality restrictions.

## Acknowledgments

Research reported in this work was supported by the National Institutes of Health under award number R01 EB028591. The content is solely the responsibility of the authors and does not necessarily represent the official views of the National Institute of Health.

